# Immunogenicity of a third dose viral-vectored COVID-19 vaccine after receiving two-dose inactivated vaccines in healthy adults

**DOI:** 10.1101/2021.09.16.21263692

**Authors:** Ritthideach Yorsaeng, Nungruthai Suntronwong, Harit Phowatthanasathian, Suvichada Assawakosri, Sitthichai Kanokudom, Thanunrat Thongmee, Preeyaporn Vichaiwattana, Chompoonut Auphimai, Lakkhana Wongsrisang, Donchida Srimuan, Thaksaporn Thatsanatorn, Sirapa Klinfueng, Natthinee Sudhinaraset, Nasamon Wanlapakorn, Yong Poovorawan

## Abstract

In June 2021, Thailand was hit by the delta variant of SARS-CoV-2 resulting in the biggest wave of COVID-19. Due to the widespread delta variant, more than 600 healthcare workers had COVID-19 despite completion of two-dose CoronaVac. The Ministry of Public Health recommended that healthcare workers received a third dose of AZD1222 to increase level of protection against SARS-CoV-2. However, immune response after the third vaccination with AZD1222 are limited. In this study, sera from those who received a booster of AZD1222 in June-July 2021 were tested for SARS-CoV-2 spike receptor-binding-domain (RBD) IgG, anti-RBD total immunoglobulins and anti-spike protein 1 (S1) IgA. The neutralizing activities in a subset of serum samples were tested against the wild type and variants of concern (B.1.1.7, B.1.617.2, and B.1.351) using an enzyme-linked immunosorbent assay-based surrogate virus neutralization test. Participants who received the booster of AZD1222 possessed higher levels of spike RBD-specific IgG, total immunoglobulins, and anti-S1 IgA than that two-dose vaccines (*p* < 0.001). They also elicited higher neutralizing activity against the wild type and all variants of concern than those in the recipients of the two-dose vaccines. This study demonstrated a high immunogenicity of the AZD1222 booster who completed the two-dose inactivated vaccines.

## 1. Introduction

Widespread vaccination against the novel severe acute respiratory syndrome coronavirus 2 (SARS-CoV-2) is crucial in mitigating the global pandemic of coronavirus disease 2019 (COVID-19). Despite the expedited development of effective vaccines, the ability to acquire sufficient vaccine supplies varies greatly between industrialized and developing countries. Beginning in February 2021, Thailand was only able to procure in large quantities the inactivated COVID-19 vaccine CoronaVac. Healthcare workers 18-59 years of age were among the first to receive two doses of CoronaVac administered 3 to 4 weeks apart. Smaller quantities of the adenovirus-vectored vaccine (AZD1222) arrived the following month, with additional doses domestically produced from licensing with AstraZeneca available several months later. Although vaccination with AZD1222 can be for adults ≥18 years, it is the only one approved for those with specific underlying comorbidities and adults ≥60 years of age. Due to the limited supply initially, vaccination between doses were 10 weeks apart.

Two doses of most COVID-19 vaccines were required to elicit an immunological response and protection against severely symptomatic disease [1-5]. Nevertheless, the emergence of SARS-CoV-2 variants with accumulated multiple mutations had raised concerns about the efficacy of the two-dose regimen [6]. Data gathered from mass vaccination in some countries have suggest that the effectiveness of mRNA [7] and AZD1222 [6] vaccines was reduced over time and varied among emerging viral variants. Therefore, many countries currently are considering the implementation of a third shot to boost the existing immune response. Ongoing clinical trials to investigate the immunogenicity, safety, and efficacy of a booster dose with the same (homologous) or different (heterologous) vaccine are underway [5, 8-9].

Since June 2021, reports of hundreds of fully vaccinated healthcare workers who were subsequently infected with the delta variants of SARS-CoV-2 have spurred the implementation of a third COVID-19 vaccine dose in Thailand, either with AZD1222 or BNT162b2 [10]. However, no data were available as to whether heterologous vaccination with an adenovirus vectored vaccine in adults who previously received two doses of an inactivated vaccine was safe and effective. To address this knowledge gap, we characterized the increase in immune response and neutralizing antibody induced by heterologous vaccination with AZD1222 in Thai healthcare workers who were previously fully vaccinated with CoronaVac.

## 2. Methods

### 2.1 Study cohort

The study flow showing participant enrollment and sample size in this study is shown in Fig. 1. The first group (n=170) received two doses of CoronaVac. The second group (n=169) received two doses of AZD1222. The third group (n=210), all healthcare workers, received two doses of CoronaVac, followed by AZD1222. An additional comparison group consisted of unvaccinated, laboratory-confirmed COVID-19 patients who have since recovered from illness as previously reported [11]. The study protocols were approved by the Institutional Review Board of the Faculty of Medicine of Chulalongkorn University (IRB numbers 192/64 and 546/64). The study is registered with the Thai Clinical Trials Registry (TCTR) with identifiers TCTR20210319003 and TCTR20210910002.

**Fig 1.**
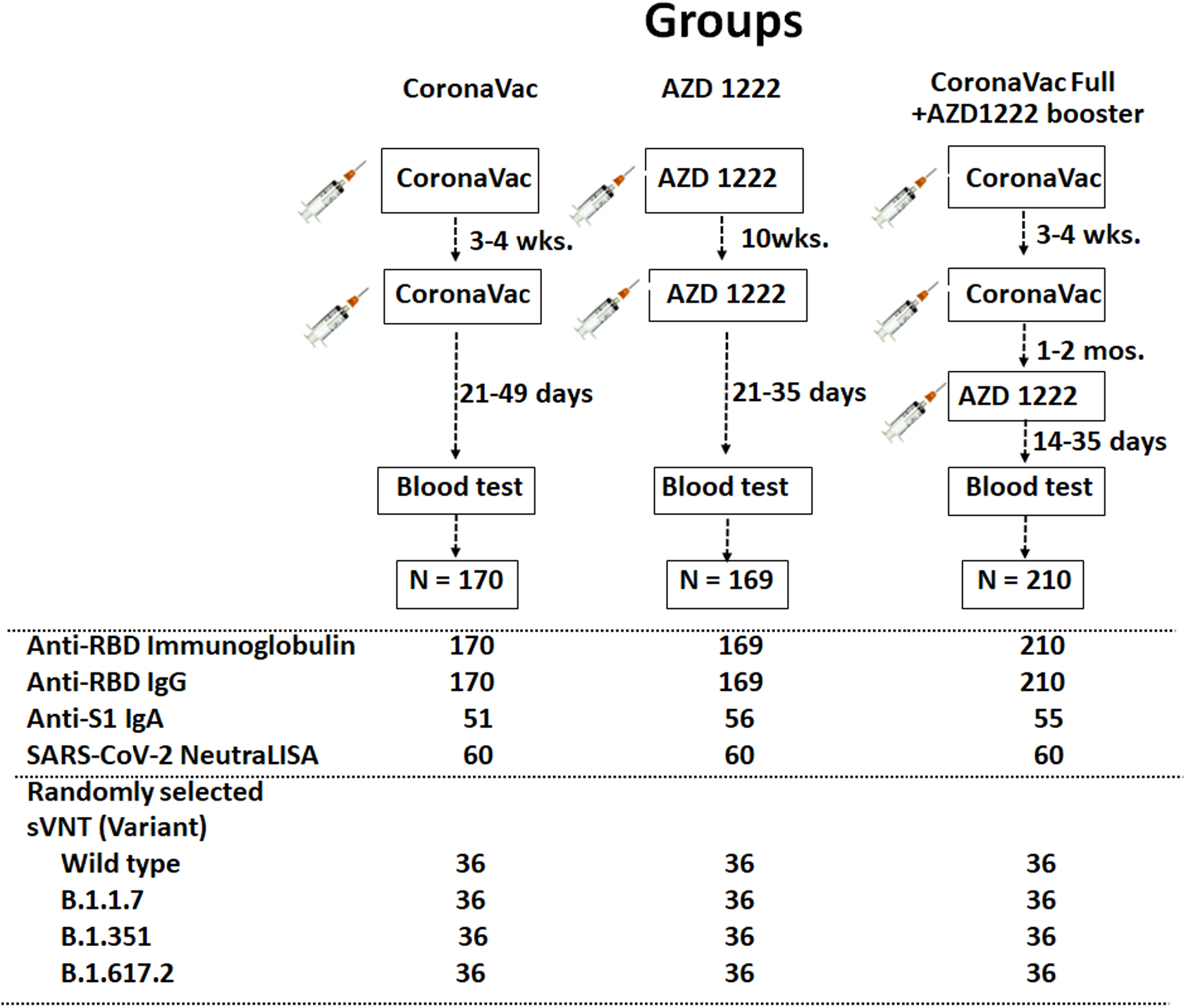
Schematic diagram of this study. Three groups of vaccinees and samples done for each test are depicted. Randomly selected samples subjected to testing listed as numbers under each group.

### 2.2 Vaccination

COVID-19 vaccines were available in Thailand beginning in March 2021. CoronaVac was approved for Thai adults ages 18-59 and was administered 21-28 days apart [12]. Since ADZ1222 was authorized for use in adults ≥18 years of age, priority was given to those ≥60 due to its limited supply in Thailand [13] and vaccination was done 10 weeks apart. Beginning in June 2021, healthcare workers who were previously fully vaccinated with CoronaVac (those who received two doses) were offered a third vaccination with AZD1222.

### 2.3 Blood samples

For the first and second group, blood samples were collected once between 2-5 weeks after the second or third vaccination. For the third group, samples were collected between 4-8 weeks after symptoms appeared or after laboratory confirmation (from testing nasopharyngeal swabs with reverse-transcription polymerase chain reaction).

### 2.4 Antibody assays

Serum samples were evaluated for total immunoglobulins specific to the receptor-binding domain (RBD) of the SARS-CoV-2 spike (S) protein using Elecsys SARS-CoV-2 S according to the manufacturer’s instruction (Roche Diagnostics, Basel, Switzerland). Values ≥0.8 U/mL was considered positive. To quantify only IgG specific to the RBD, we utilized SARS-CoV-2 IgG II Quant assay (Abbott Diagnostics, Abbott Park, Ill) according to the manufacturer’s instruction. Values ≥50 arbitrary U/mL (AU/mL) was considered positive. Multiplying the numerical AU/mL by 0.142 converts it to binding antibody units per milliliter (BAU/mL). A subset of samples was evaluated for anti-S1 IgA using an enzyme-linked immunosorbent assay (ELISA) according to the manufacturer’s instruction (Euroimmun, Lübeck, Germany). The results were derived from the ratio in optical density (OD) obtained from samples and the calibrator. Maximum cut-off ratio (OD/CO) was 9; results >9 was recorded as 9.

A subset of samples were also evaluated for neutralizing activity against the SARS-CoV-2 wild-type and variants of concern B.1.1.7 (alpha), B.1.351 (beta), and B.1.617.2 (delta) using an ELISA-based surrogate virus neutralization test (sVNT). NeutraLISA (Euroimmun, Lübeck, Germany) was used to evaluate the wild-type strain only. Results ≥35% inhibition was considered positive. Additionally, cPass SARS-CoV-2 neutralizing antibody detection kit (GenScript, Piscataway, NJ) was used for all strains. The recombinant RBD from B.1.1.7 (containing N501Y), B.1.351 (containing N501Y, E484K, and K417N), and B.1.617.2 (containing L452R and T478K) were also used with this kit. Briefly, the serum samples were diluted 1:10 with buffer and incubated with RBD conjugated to horseradish peroxidase for 30 min. at 37°C. Next, 100 µL of the sample mixture was added to a capture plate pre-coated with human angiotensin-converting enzyme 2 (ACE2) and incubated for 15 min at 37°C. After washing, 100 µL of TMB chromogen solution was added and the plate incubated in the dark for 15 min at room temperature. After the addition of 50 µL stop solution, samples were read at 450 nm. The ability of a serum to inhibit binding between RBD and ACE2 was calculated as percentage as follows: 1 – (average OD of sample/average OD of negative control), multiplied by 100. Inhibitions ≥30% was considered positive.

### 2.5 Statistical analysis

Baseline characteristics were reported as mean and standard deviations (SD), or median and range. Antibody levels were presented as geometric mean titers (GMT) with 95% confidence interval (CI), or median with interquartile range when appropriate. The difference in baseline characteristics such as sex and age were calculated by Chi-squared test and *t*-test, respectively. The difference in antibody titers, OD/CO, and percent inhibition between groups was calculated using ANOVA or the Kruskal–Wallis test. Statistical analysis was done using Prism 8.0 (GraphPad, San Diego, CA). A *p* value <0.05 was considered statistically significant.

## 3. Results

### 3.1 Demographic data of vaccinees

The fully vaccinated AZD1222 group was older (mean age 47.9 years). There were significantly more women in the fully vaccinated CoronaVac + AZD1222 group (Table 1).

**Table 1.**
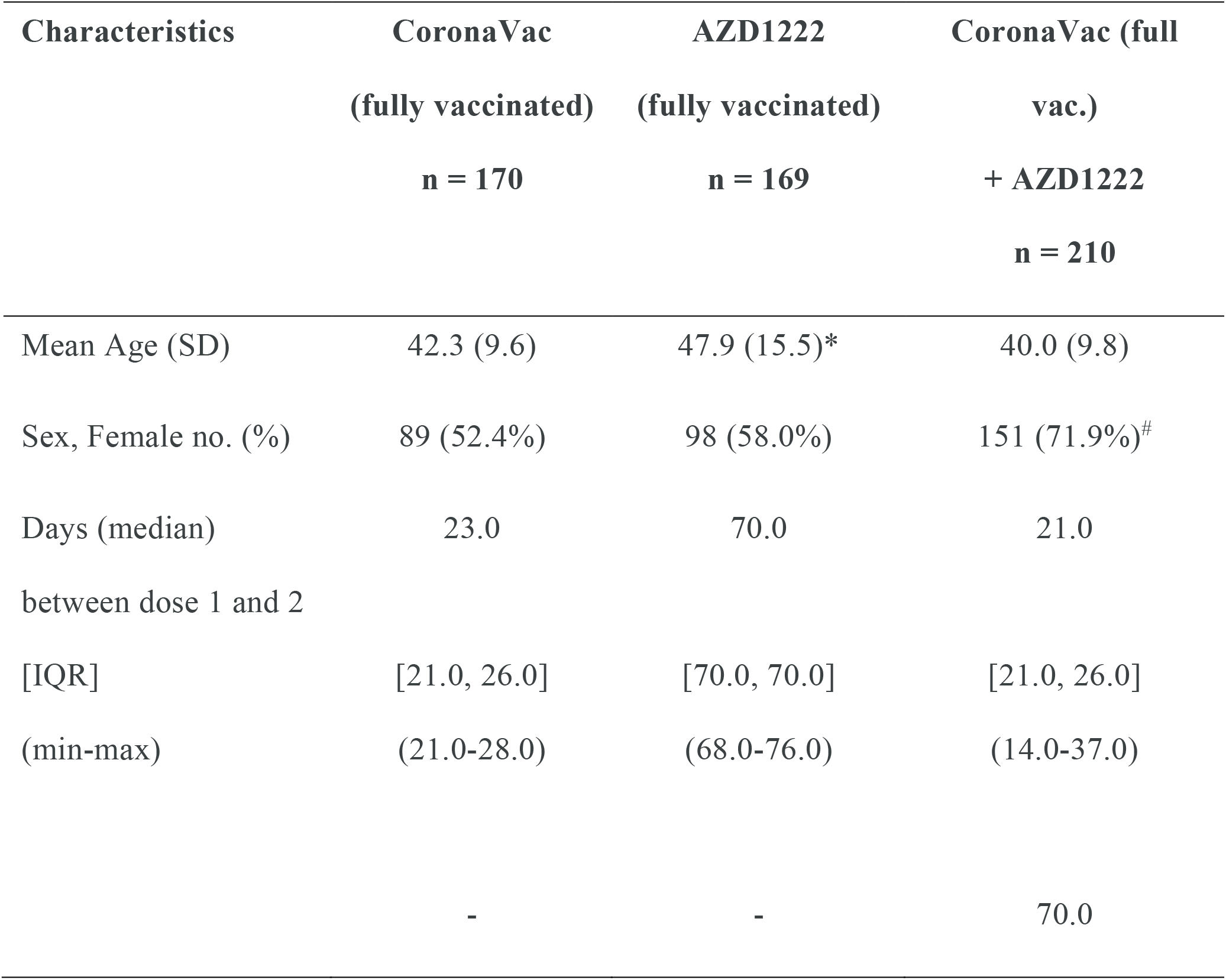
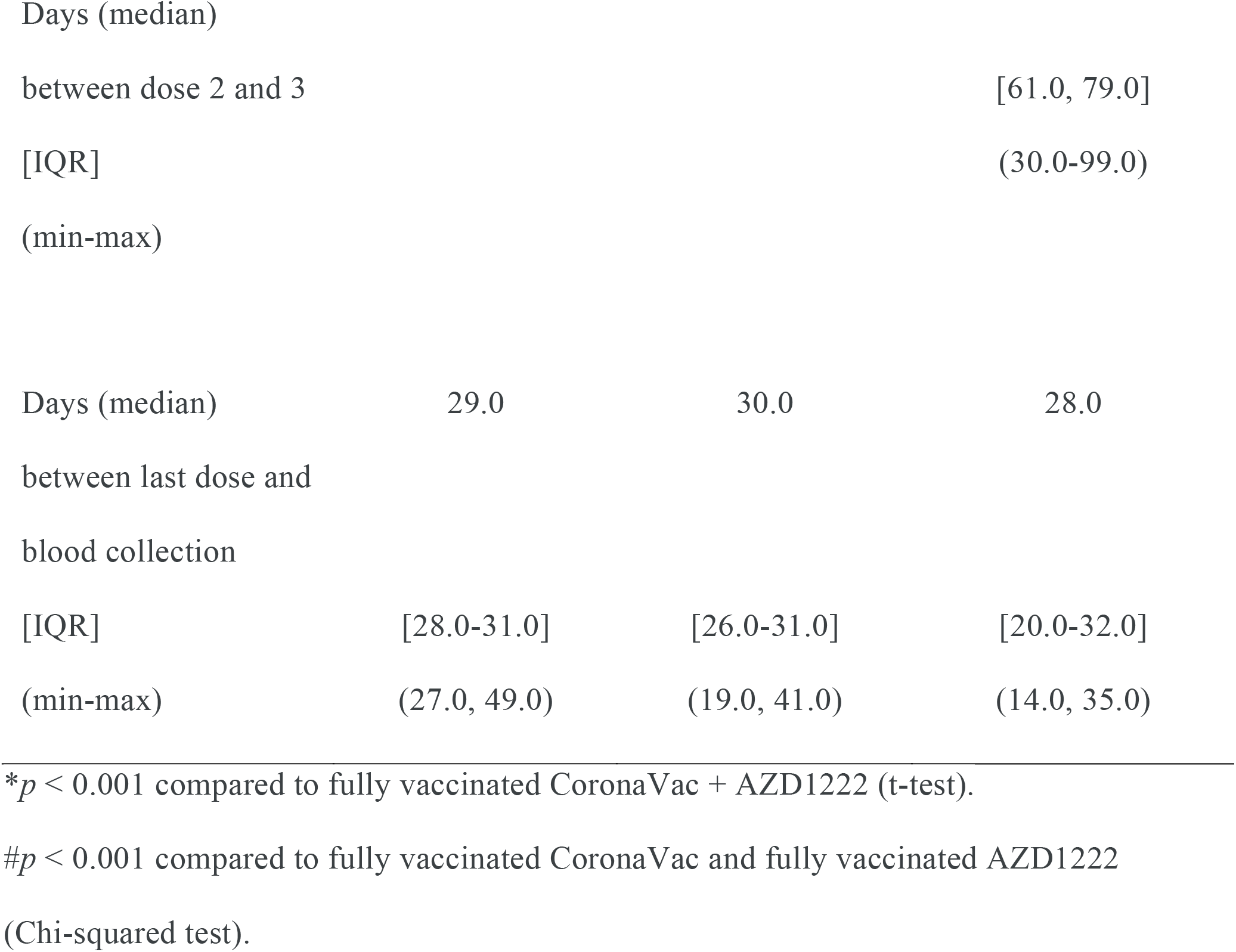
Characteristics of the vaccinees in this study

### 3.2 sRBD-specific SARS-CoV-2 antibodies

We first evaluated overall seroconversion in vaccinated individuals and recovered COVID-19 patients. The GMT of total anti-RBD immunoglobulins among vaccinees fully vaccinated with CoronaVac was comparable to that of unvaccinated convalescent patients (<100 U/mL) (*p* = 0.7608) (Fig. 2a.). The GMT of fully vaccinated CoronaVac + AZD1222 (7,947 U/mL) was nine-fold greater than that of fully vaccinated AZD1222. When we determined only the contribution of anti-RBD IgG, the GMT among convalescent patients (28.1 BAU/mL) was lower than that of fully vaccinated CoronaVac group (128 BAU/mL) (*p* = 0.0375). In contrast, the third booster dose with AZD1222 induced the most antibody response (1,492 BAU/mL) compared to other groups (Fig. 2b.). Serum anti-S1 IgA was also highest in this group (Fig. 2c.). The presence of serum anti-S1 IgA was observed only after a third vaccination and in convalescent patients. In addition, the sVNT assay, which was based on antibody-mediated inhibition of ACE2 and S protein interaction, showed that the fully vaccinated CoronaVac + AZD1222 recipients had higher neutralizing activities (demonstrated as percent inhibition) against the original Wuhan (wild-type) strain compare to fully vaccinated CoronaVac and fully vaccinated AZD1222 groups (Fig. 2d.).

**Fig 2.**
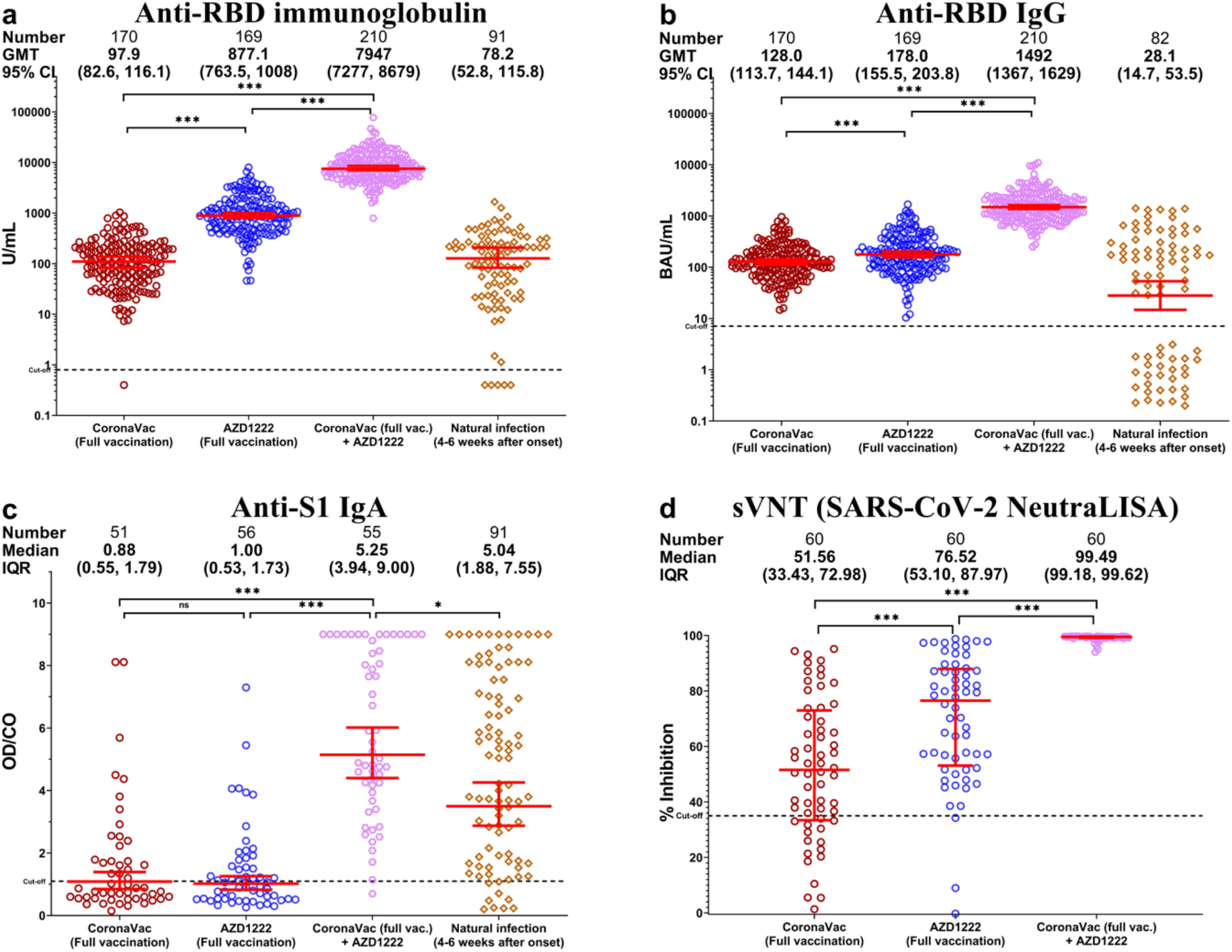
Immunoassays for SARS-CoV-2 in serum samples from fully vaccinated and unvaccinated convalescent COVID-19 patients. (a) Total anti-RBD immunoglobulins. (b) Anti-RBD IgG. (c) Anti-S1 IgA. (d) Surrogate virus neutralization test (sVNT). Geometric mean titer (GMT) with 95% confidence intervals are shown as red lines within each group. * denotes *p* < 0.05 and *** denotes *p* ≤ 0.001.

### 3.3 Neutralizing antibodies against SARS-CoV-2 variants

Although serum samples from the fully vaccinated AZD1222 group was able to better inhibit wild-type SARS-CoV-2 and variants than those from the fully vaccinated CoronaVac (*p* < 0.01), the fully vaccinated CoronaVac + AZD1222 group demonstrated higher neutralizing antibodies compared to two doses of either vaccine (*p* < 0.0001) (Fig. 3a-d.).

**Fig 3.**
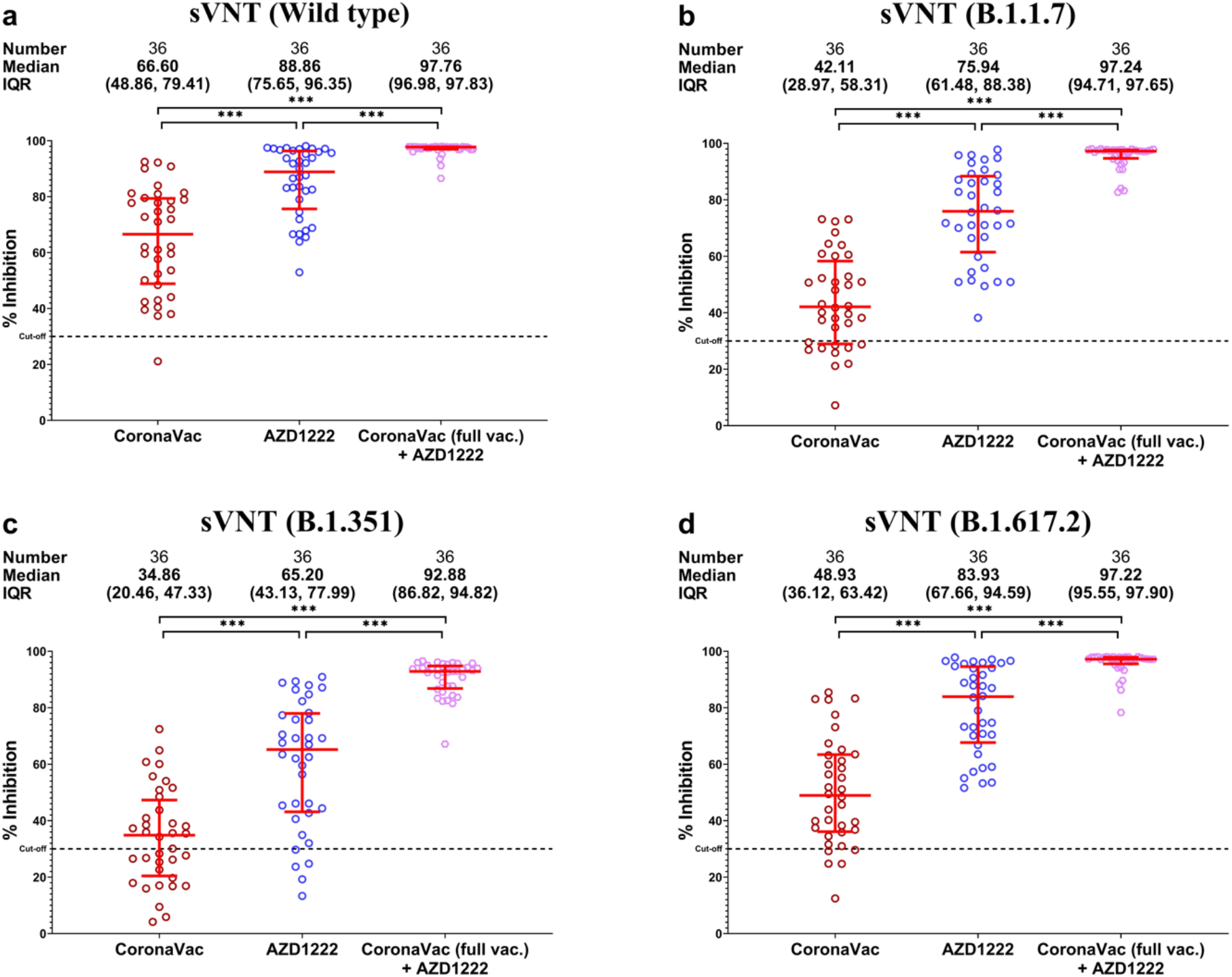
Surrogate virus neutralization test against SARS-CoV-2 wild-type and variants. (a) Wild-type. (b) B.1.1.7. (c) B.1.351. (d) B.1.617.2. Percent inhibition is shown with mean and IQR (red lines). *** denotes *p* ≤ 0.001.

Comparison of neutralizing activities within the same participants against the wild-type and variant strains showed that all vaccinated groups elicited higher neutralizing activities in the following order: wild type > B.1.617.2 > B.1.1.7 > B.1.351 (Fig. 4a-c.). The fully vaccinated CoronaVac group showed the lowest inhibition against B.1.351 (Fig. 3c.).

**Fig 4.**
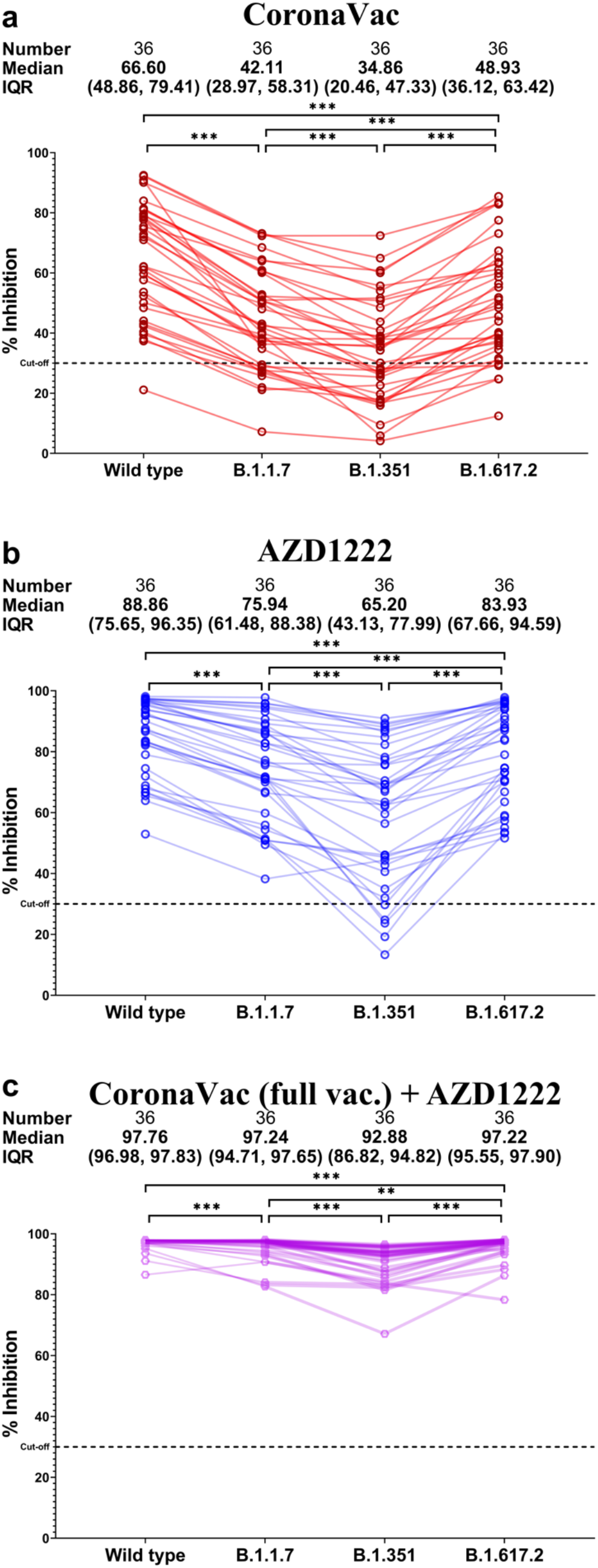
Comparison between neutralizing activities against variants relative to the wild type in serum samples obtained from (a) two-dose CoronaVac/CoronaVac, (b) two-dose AZD1222/AZD1222, and (c) two-dose CoronaVac + one-dose AZD1222 vaccinees. *** denotes *p* ≤ 0.001. ** denotes *p* ≤ 0.01.

## 4. Discussion

Recent data suggest that the immunity elicited by two-dose vaccination against COVID-19 waned over time resulting in reduced protection against wild type SARS-CoV-2 and variants of concerns [14], calling into question whether a third dose would be advantageous or potentially required for long-term protection. This study showed that participants who received the third dose AZD1222 vaccine after completion of the two-dose CoronaVac vaccines possessed higher levels of spike RBD-specific IgG, total immunoglobulin, and anti-S1 IgA than that of two-dose CoronaVac and AZD1222 vaccines. In addition, sera samples from booster dose vaccine recipients elicited higher neutralizing activity against the wild type and all variants of concern than those in the recipients of the two-dose CoronaVac and AZD1222 vaccines.

These results follow similar trends expressed in research on the immunogenicity of a homologous three-dose regimen of the CoronaVac vaccine [8, 15], AZD1222 vaccine or mRNA-1273 vaccine [16]. Commonly, it was shown that a third dose evokes a higher immunological response surpassing levels post-second dose of the primary vaccines as a booster effect. In addition, mouse models using the heterologous vaccine regimen have shown that the three-dose vaccine schedule can promote higher humoral and cellular responses than that induced by the same three-dose regimen [17]. Furthermore, sVNT experiments presented significantly showed higher post-third dose inhibition levels of neutralizing activities against the wild type and variants of concerns in the third dose vaccine recipients, which is consistent with that of a high immunoglobulin titer.

Aside from heightened immunoglobulin titers and sVNT inhibition percentages, a notable discrepancy of immunoglobulin subtype A was observed. Serum anti-spike1 protein-specific IgA OD/CO ratios significantly increased following an AZD1222 vaccine as a third booster, compared to after two-dose homologous CoronaVac and AZD1222 regimen. The presence of serum spike-specific IgA was observed only after a third vaccination and natural infection. IgA is a potent immunoglobulin [18] found mainly in mucosal regions and is fundamental for inhibiting respiratory viruses from epithelial cell attachment [19]. Despite differing mechanistic characteristics, our results highlighting an absence of serum anti-spike specific IgA after a two-dose homologous CoronaVac and AZD1222 schedule. In contrast, some two-dose regimens can elicit serum IgA responses despite the absence of mucosal IgA responses. Limited data is available on the clinical benefit of serum IgA in protection, however, a recent in vitro experimental study has shown that serum IgA contributed to the neutralization of the SARS-CoV-2 [18, 20, 21].

Our study had a few noteworthy limitations. First, only immunological data was collected and therefore this investigation lacks information regarding the efficacy of a heterologous third AZD1222 vaccination. Second, demographic data collected highlighted a gender disparity with a large majority of female participants in the booster group. Additionally, a difference in the average age between groups was noted between homologous and the three-dose regimen. Lastly, a cell-mediated response was not examined. Only a humoral response was investigated, accounting for a portion of an immunological response to vaccination.

In conclusion, a three-dose heterologous regimen, two initial CoronaVac followed with a third AZD1222 vaccine, indicated a strong immunological response. Additional research into age-related efficacy, vaccination timing, and cell-mediated response, and effectiveness data should be considered for a comprehensive understanding of the benefit of this vaccine schedule. Further investigations into the vaccine-induced protection against SARS-CoV-2 emerging variants and their impact on this vaccination regimen’s effectiveness are to be further investigated.

## Data Availability

The availability of all data referred to in the manuscript.

## Author Contributions

Conceptualization, R.Y., N.S., N.S., N.W. and Y.P.; data collection, D.S. and T.T.; formal analysis, R.Y., N.S., N.W. and Y.P.; methodology, N.S., S.A., S.K., T.T., P.V., C.A. and L.W.; project administration, Y.P.; writing-original draft, R.Y., H.P., N.W. and Y.P.; writing-review and editing, N.W., N.S. and Y.P.; All authors have read and agreed to the published version of the manuscript.

## Institutional Review Board Statement

The study protocols were approved by the Institutional Review Board of the Faculty of Medicine of Chulalongkorn University.

## Informed Consent Statement

This study was carried out following the principles expressed in the Declaration of Helsinki. The written inform consent was obtained from the CoronaVac, AZD1222 and natural infection groups. Patient consent in the CoronaVac Full + AZD1222 group was waived due to the fact, that the datasets used in this study were anonymous and prevent the identification of any individual study subject by the research team at any stage of the study.

## Data Availability Statement

The datasets generated during and/or analyzed during the current study are available from the corresponding author on reasonable request.

## Acknowledgements

We would like to thank all the staffs of the Center of Excellence in Clinical Virology and all the participants for helping and supporting in this project.

