## Supplementary Table 1 for "Immunogenicity of a third dose viral-vectored COVID-19 vaccine after receiving two-dose inactivated vaccines in healthy adults"

**Supplementary Table 1.** Total anti-RBD immunoglobulins, Anti-RBD IgG, Anti-S1 IgA, Surrogate virus neutralization test against Wild-type, B.1.1.7, B.1.351 and B.1.617.2 in the vaccinees in this study.

| Results | **CoronaVac/CoronaVac (n = 170)** | **AZD1222/AZD1222 (n = 169)** | **CoronaVac (full vac.) + AZD1222 (n = 210)** |
|---|---|---|---|
| Immunoglobulin anti-RBD (U/mL) GMT (95% CI) | 97.94 (82.59, 116.1) | 877.1 (763.5, 1008) | 7947 (7277, 8679) |
| IgG anti-RBD (BAU/mL) GMT (95% CI) | 128.0 (113.7, 144.1) | 178.0 (155.5, 203.8) | 1492 (1367, 1629) |
| IgA anti-S1 (OD/CO) Median [IQR] (min-max) | 0.88 [0.55, 1.79] (0.15-8.11) | 1.00 [0.53, 1.73] (0.26-7.30) | 5.25 [3.94, 9.00] (0.70-9.00) |
| sVNT (Wild-type, Euroimmun) (% Inhibition) [Median (IQR)] | 51.56 [33.43, 72.98] (1.36-95.17) | 76.52 [53.10, 87.97] (-0.27-98.76) | 99.49 [99.18, 99.62] (94.03-99.79) |
| sVNT (Wild-type, | 66.60 | 88.86 | 97.76 |

| | | | |
|---|---|---|---|
| GenScript) (% Inhibition) [Median (IQR)] | [48.86, 79.41] (21.12-92.50) | [75.65, 96.35] (52.94-98.12) | [96.98, 97.83] (86.56-97.99) |
| sVNT (B.1.1.7, GenScript) (% Inhibition) [Median (IQR)] | 42.11 [28.97, 58.31] (7.21-73.15) | 75.94 [61.48, 88.38] (38.23-97.80) | 97.24 [94.71, 97.65] (82.67-98.07) |
| sVNT (B.1.351, GenScript) (% Inhibition) [Median (IQR)] | 34.86 [20.46, 47.33] (4.15-72.43) | 65.20 [43.13, 77.99] (13.33-90.92) | 92.88 [86.82, 94.82] (67.14-96.56) |
| sVNT (B.1.617.2, GenScript) (% Inhibition) [Median (IQR)] | 48.93 [36.12, 63.42] (12.46-85.45) | 83.93 [67.66, 94.59] (51.62-97.83) | 97.22 [95.55, 97.90] (78.33-98.09) |
